# Different perspectives on Quality in clinical education of physical therapy students – protocol of a scoping review

**DOI:** 10.1101/2023.10.20.23297304

**Authors:** Matthias M Walter, Slavko Rogan, Alexander P Schurz, Evert Zinzen

## Abstract

**Background:** Clinical education (CE) plays a crucial role in physical therapy education, yet there is a notable absence of established quality characteristics for its implementation.

**Objectives:** This scoping review aims to elucidate how various stakeholders define and describe the quality of CE in higher education for physical therapy students. Additionally, it seeks to identify commonalities and distinctions in the application of the term “quality” in the context of CE.

**Methods:** Peer-reviewed studies encompassing physical therapy students, clinical instructors, lecturers in physical therapy education, physical therapy educational sites, and supervising physical therapists in internships across all clinical fields will be included in the review. A comprehensive search strategy will be employed, utilizing multiple electronic databases, including MEDLINE, EMBASE, the Cochrane Library, ERIC, Education Research Complete, Education Database, and CINAHL. Eligibility screening will be independently conducted by two reviewers. Data extraction will be presented in a tabular or graphical format, aligning with the review’s objectives.

**Discussion:** Insights gleaned from this study hold the potential to inform targeted interventions and improvements in CE, ultimately enhancing the learning outcomes and satisfaction of physical therapy students. This endeavor seeks to bridge the existing gap in defining and achieving quality in clinical education within the realm of physical therapy higher education.

## Introduction

Clinical education (CE) is a pivotal component of physical therapy education, constituting up to one third of the curriculum (1,2). It entails exposing students to genuine clinical practice, which differs from the theoretical aspects typically presented in classroom settings. During CE, the foundation for developing therapeutic competencies is formed (2). Given its substantial role in shaping future practitioners, maintaining high-quality CE experiences is of paramount importance to ensure the competence and excellence of future physical therapists (3).

At the level of physical therapy educational sites Directors of CE aim to control and increase the quality of CE. However, currently, there are no quality characteristics for its implementation. The one universal requirement, as mandated by the accreditation criterion, is that physical therapy education programs must demonstrate their graduates’ ability to effectively manage patients and clients across a wide range of practice settings and throughout different stages of life and care continuum (4).

Physical therapy educational sites operate with varying personnel and financial resources, which impact the learning experience of students and leads to significant heterogeneity in quality of education (5). Furthermore, human resources, materials as well as other factors which could influence CE experiences (CEEs) are mostly managed by clinical practices and thus, are outside of the control of Directors of CE. Nevertheless, it is the responsibility of Directors of CE to provide high-quality CEEs (6).

In the realm of CE for physical therapists, numerous stakeholders play pivotal roles in shaping the learning experience. These key stakeholders include physical therapy students, educators, clinical instructors, supervising physical therapists within healthcare facilities, and physical therapy educational sites. Each of these stakeholders contributes unique perspectives on what constitutes quality in CE. There are many factors within the structures and processes of CE, and they may interact with one another to contribute to various outcomes. Thus, the direction of students’ learning and satisfaction is a collaboration of these stakeholders (7).

The intricacy of assessing the quality of CE is highlighted by a systematic review, which yielded insufficient evidence to ascertain the best practices for CE in physical therapy in general (8). A recently developed tool by (9) seems to assess quality in clinical education. However, this tool should be answered by Directors of CE and heads of departments and is therefore missing the evaluation of quality from a physical therapy student perspective.

Evaluating stakeholder satisfaction has the potential to serve as a valuable resource in ongoing endeavors to enhance quality in CE. However, to develop a tool to assess satisfaction and quality in CE, it should be clarified what defines quality in CE from the different perspectives of stakeholders. Therefore, the primary aim of this scoping review is to map how different stakeholders define and describe quality of CE in higher education of physical therapy students. Secondary aim is to outline the similarities and differences in the use of the term quality in CE.

## Methods

This scoping review follows the Joanna Briggs Institute Evidence recommendations (10–13) and the Preferred Reporting Items for Systematic Review and Meta-Analysis Extension for Scoping Reviews checklist (PRISMA-ScR) (14).

### Research question of this scoping review

This scoping review aims to answer the following research questions:

1. How do different stakeholders (students, clinical instructors, supervising PTs, PT educational sites) define and describe quality of CE in higher education of physiotherapy students?
2. What are the similarities and differences in the use of the term quality in CE?

### Protocol and registration

This protocol was preregistered in the International Database of Education Systematic Reviews under the registration number [IDESR000098]. Any significant updates or changes to this protocol will be documented in the same registration entry to ensure transparency and traceability throughout the research process.

### Eligibility criteria

The current scoping review will consider peer-reviewed studies that include physical therapy students, clinical instructors and/or lecturers in physical therapy education, physical therapy educational sites as well as supervising physical therapists in internships from all clinical fields. Only English language articles will be included, and we will apply no time limit to the search strategy. Studies from any geographical and clinical setting will be eligible for inclusion. Investigations focusing on any kind of quality (e.g., satisfaction, learning experiences) in clinical education will be included.

Exclusion criteria include other healthcare professions than physical therapy, study protocols and abstracts, conference proceedings, and dissertations.

### Information source

The search strategy will encompass multiple electronic databases, including MEDLINE (accessed via Ovid), EMBASE, the Cochrane Library, ERIC, Education Research Complete, Education Database and CINAHL (accessed via Ebsco).

### Search

We will use the PCC (Population (or participants)/Concept/Context) framework which is recommended by the Joanna Briggs Institute (11). A comprehensive search strategy will be conducted to identify relevant studies. The initial search strategy will focus on the electronic database MEDLINE (PubMed). The researchers will analyze text words in article titles, abstracts, and index terms related to the research topic. The identified keywords and index terms will be customized for each information source and combined using Boolean operators, forming the final search strategy. In addition to electronic databases, the researchers will explore the reference lists of included studies to uncover further relevant studies. The process of identifying relevant studies will be completed by the end of October 2023. Table 1 represents the pilot search strategy.

**Table 1.**
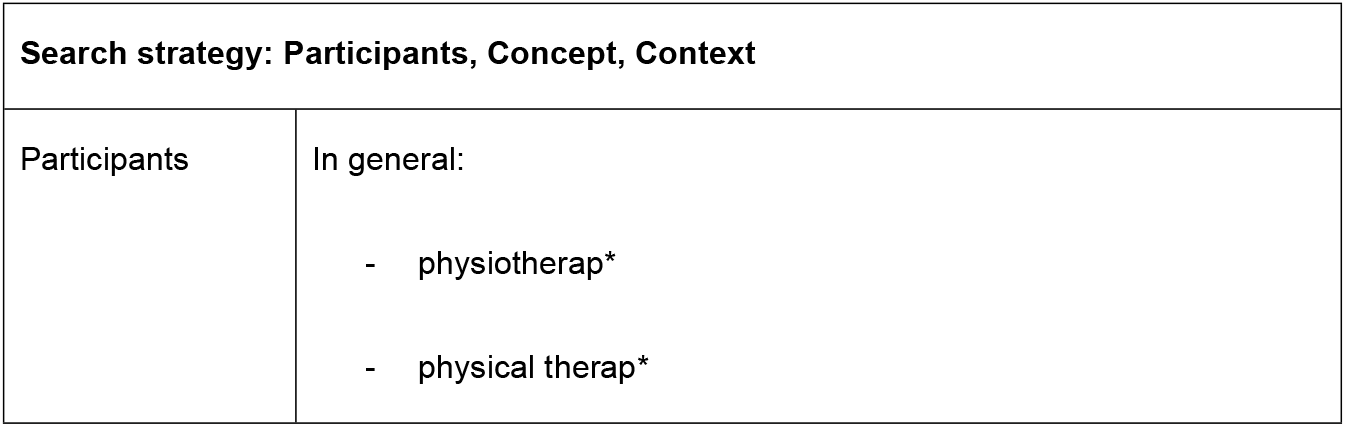
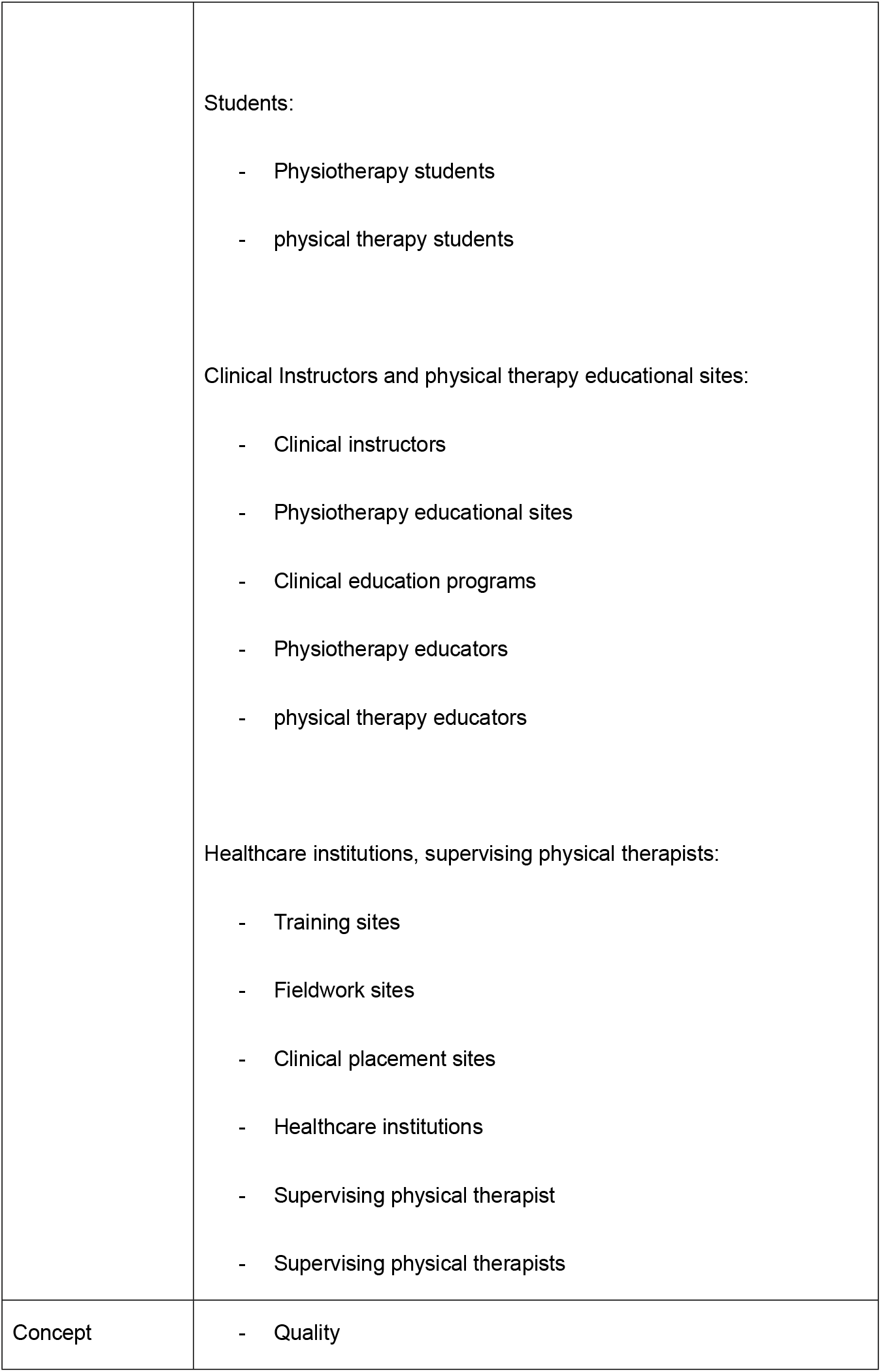
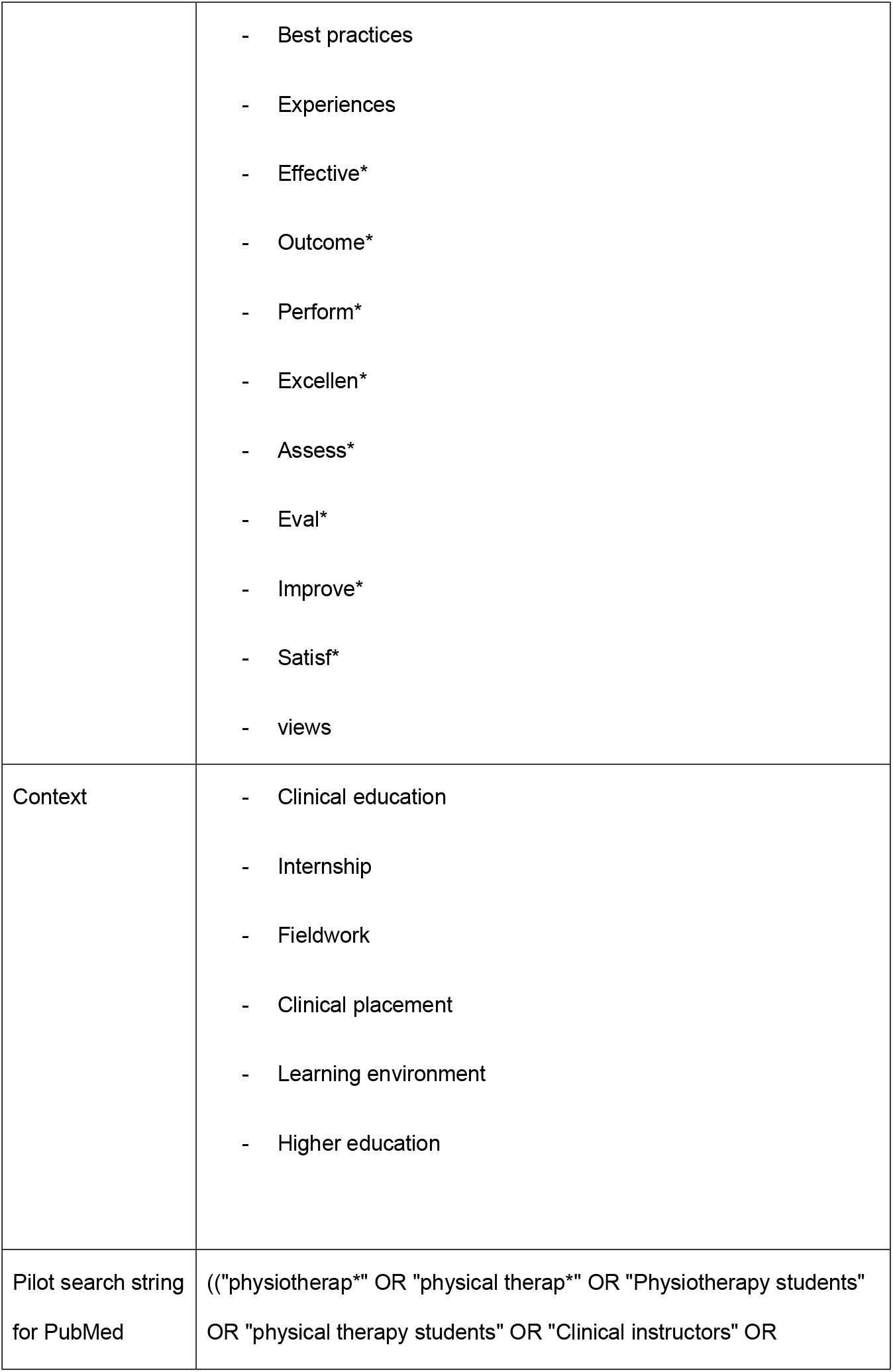
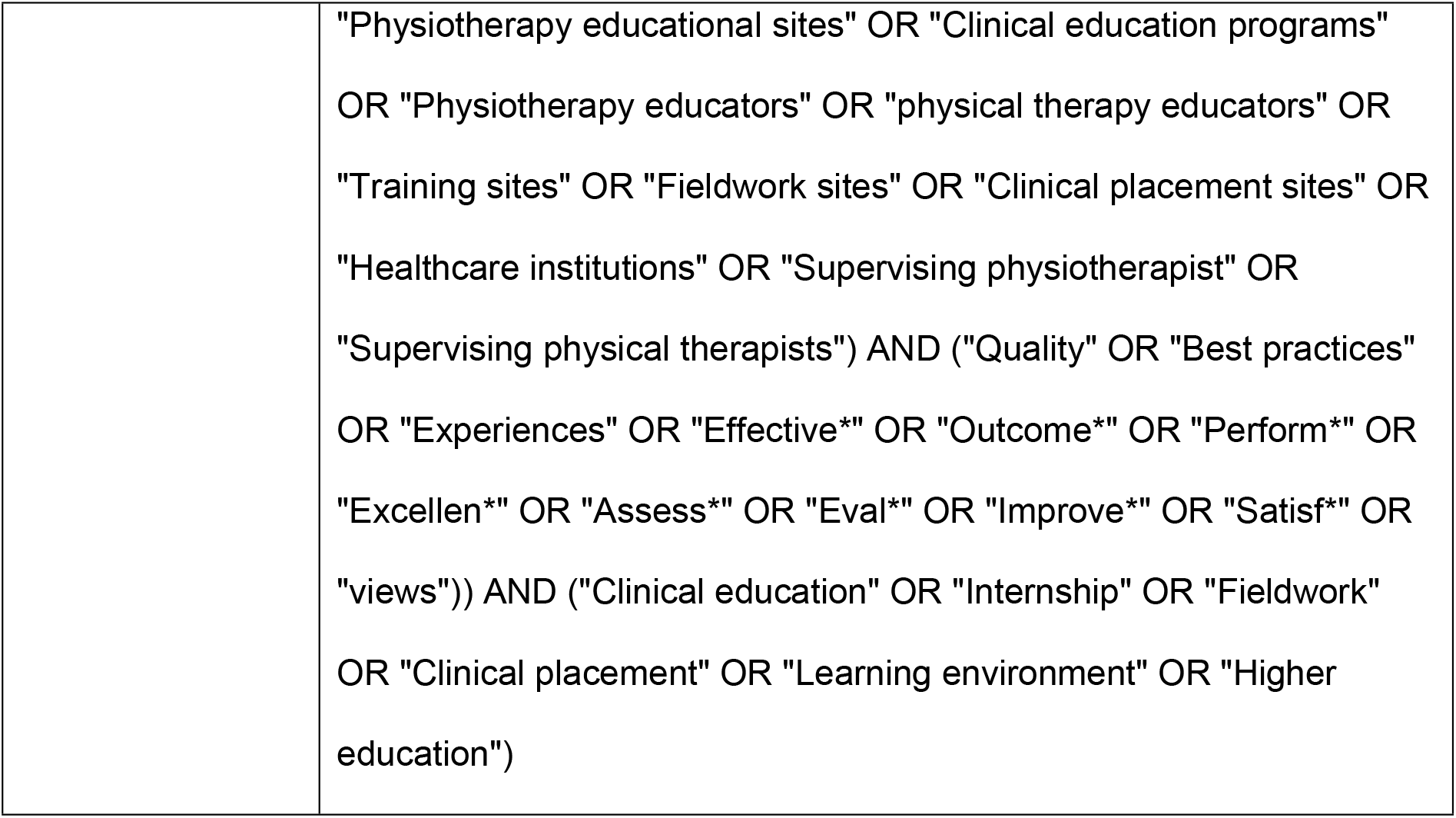
Search strategy.

#### Selection of sources of evidence

First, two reviewers (MW, AS) will independently screen title and abstract for eligibility criteria. Any articles deemed relevant by both reviewers will be included in the full-text review. If the relevance of a study was unclear from the abstract, then the full article was ordered. Second, two investigators (MW, AS) will conduct an independent assessment of the full-text articles to ascertain their compliance with the predefined eligibility criteria. To gauge the inter-rater agreement, Cohen’s κ statistic will be computed for both the initial title and abstract review stage, as well as for the subsequent full article review stage. In cases where any discrepancies arise during the evaluation of full-text articles, a reevaluation of these discordant articles will be performed. Any persisting disagreements regarding the eligibility of studies will be diligently resolved through constructive discussions with a third reviewer (SR). The goal of these discussions will be to achieve complete consensus among all reviewers regarding the inclusion of studies. We will use Rayyan for the screening process (15).

#### Data charting process

Data from the articles included will be extracted by two independent reviewers (MW, AS) into a modified version of the JBI data extraction table (16). To identify any necessary adjustments to the data extraction table, we will conduct a pilot test of the data extraction table using three full texts before proceeding with data extraction. In case of any disagreements a third reviewer (SR) will be involved to find consensus. Where required, authors will be contacted to request missing or additional data. Due to the primary objective of scoping reviews to comprehensively map and summarize the existing evidence on a topic, rather than synthesizing the findings to address a specific clinical question, the evaluation of methodological limitations or the risk of bias in the included evidence is usually not undertaken, unless explicitly demanded by the specific aims of the scoping review (11,14,17). Therefore, we did not conduct a risk of bias assessment.

### Data items

The extracted data will include details about the authors, healthcare settings, Characteristics of included publications (Contexts & Participants), Location(s) of data collection, other relevant socio demographic information, publication year, study design, study methods, results and authors’ interpretation or conclusion of significance to the scoping review questions and objectives. For a detailed prescription of the included details see supplementary file 1.

### Synthesis of results

The data extracted from the included studies will be subjected to a two-step analysis: descriptive and thematic. In the first step, the extracted data will be presented in a tabular or graphical format, in line with the objectives of this scoping review. These tables and/or charts will outline the diverse perspectives on quality in CE, as described by different stakeholders. The thematic analysis will identify key themes related to the definitions and descriptions of quality. A narrative summary will be provided to interpret the results and demonstrate how they relate to the review’s overall objectives and research questions.

## Limitation

One notable limitation is the potential for publication bias. This study relies on published works, possibly excluding unpublished research, gray literature, or studies not accessible through the selected databases. This could lead to an incomplete representation of the available evidence, potentially affecting the results.

Furthermore, the primary objective of this scoping review is to explore quality perspectives from a diverse group of stakeholders, including students, educators, clinical instructors, and supervising physical therapists. This diversity may result in varying interpretations and definitions of quality. Consequently, synthesizing findings across these different perspectives may prove challenging and may introduce subjectivity.

Moreover, this study encompasses perspectives from different countries and educational systems, each with its unique context and approach to physical therapy education. These variations may introduce differences in the understanding and measurement of quality. As a result, the findings may not be universally applicable to other countries or regions.

Lastly, it’s important to note that this scoping review does not assess the methodological limitations or risk of bias in the individual studies included. Variations in the quality of these studies may impact the overall findings and interpretations, potentially affecting the robustness of the conclusions drawn.

## Discussion

We plan to conduct a scoping review using the scoping review methodological framework by the Joanna Briggs Institute Evidence recommendations to ensure the replicability of our study and to reinforce our methodology (10–13). This scoping review would like to represent factors that influence various aspects related to CE in physical therapy education. The findings of the scoping review proposed should be made available within the broadest possible audience. They should understand the different perspectives of quality in CE to help educational institutions and clinical sites identify areas for improvement. Insights gained from this study may lead to targeted interventions and enhancements of CEEs, ultimately benefiting physical therapy students’ learning outcomes and satisfaction.

The findings of this review could inform policymakers and regulatory bodies involved in physical therapy education. By identifying different stakeholders’ perspectives on quality in CE, the study may contribute to the development of standardized procedures and guidelines for implementing and assessing CE programs.

This scoping review’s results may facilitate better collaboration and communication among key stakeholders, including students, educators, clinical instructors, and supervising physical therapists. By recognizing and addressing differences in how each group defines and describes quality, stakeholders can work together to create more cohesive and effective CEEs. This could contribute to the development of tools for assessing stakeholder satisfaction in CE. By understanding the elements that different stakeholders consider essential for quality, researchers and educators can design assessment instruments that capture diverse perspectives comprehensively.

Furthermore, the scoping review may also identify research gaps and areas for further investigation related to CE quality. By highlighting aspects that have received less attention or exploration, the study may inspire future research endeavors aimed at addressing unmet needs in the field.

Overall, this scoping review has the potential to make a significant contribution to the field of physical therapy education by shedding light on different perspectives of quality in CE and its impact on students’ experiences and outcomes.

## Data Availability

No datasets were generated or analysed during the current study. All relevant data from this study will be made available upon study completion.

## Ethics and dissemination

This scoping aims to provide a comprehensive overview of how various stakeholders, including students, clinical instructors, supervising PTs, and PT educational sites, define and describe the concept of quality in CE for higher education of physical therapy students. Additionally, the review seeks to identify similarities and differences in the usage of the term “quality” within the context of CE. Due to the nature of a scoping review and its secondary data analysis ethical review is not required. The results of this scoping review will be disseminated through a peer-reviewed publication to ensure the rigor and credibility of the findings.

## Contribution of Authors

MW and SR originated the concept, while EZ provided supervision and substantial input. AS offered substantial input. MW and SR crafted the search strategy, and all authors have reached a consensus on the current protocol for this scoping review.

## Acknowledgements

We have no acknowledgments to make as there was no external funding or support for our research.

## Declaration of interest

All authors declare no conflicts of interest.

## Sources of support

There are no sources of support to report.

## Notes

### Competing Interest Statement

The authors have declared no competing interest.

### Funding Statement

The author(s) received no specific funding for this work.

